# Sex-Modulated Coupling Between Genetically-Regulated Gene Expression and Functional Network Connectivity in Schizophrenia

**DOI:** 10.64898/2026.09.09.26362641

**Authors:** Guozheng Feng, Pablo Andrés-Camazón, Zening Fu, Vince D. Calhoun, Armin Iraji, Jiayu Chen

## Abstract

Schizophrenia is associated with distributed genetic, molecular, and functional brain network abnormalities, where sex differences have been well documented. However, whether sex difference exists in genetically regulated molecular variation, and how sex impacts the relationship between regulatory liability and large-scale functional dysconnectivity in schizophrenia, remain largely unclear. Here, we integrated brain eQTL-predicted gene-expression features with resting-state functional network connectivity (FNC) using an independent multimodal decomposition framework. In 367 participants from FBIRN and COBRE, gene-expression and FNC features were independently decomposed into latent components and screened for sex differences and schizophrenia associations. We identified a schizophrenia-associated gene-expression component that also showed a significant sex difference and was enriched in cerebellar tissues and biological processes related to vesicle and membrane organization and intracellular trafficking. This gene-expression component was significantly associated with a schizophrenia-associated FNC component involving cortical, subcortical, and cerebellar systems. Particularly, the gene-expression and FNC components also presented a significant sex-by-gene interaction characterized by a stronger negative association in males and a weaker positive association in females. In UK Biobank, the schizophrenia-associated gene-expression effect replicated in 2,273 individuals with schizophrenia and matched controls, while the multimodal association and its sex-dependent interaction also replicated in 4,206 participants with matched genetic and imaging data. These findings suggest that schizophrenia-related regulatory and functional-network abnormalities may converge across biological scales and that their coupling may be moderated by sex.

## INTRODUCTION

Schizophrenia is a highly heritable psychiatric disorder with a complex polygenic architecture. Large-scale genome-wide association studies have identified hundreds of risk loci enriched near genes expressed in excitatory and inhibitory neurons and involved in synaptic organization, differentiation and transmission^1^. Additional evidence from functional annotation studies suggests that the effects of common schizophrenia risk variants are more frequently mediated through regulatory mechanisms affecting gene expression than through alterations to protein-coding sequence^2–5^. Consistent with this, brain expression quantitative trait loci (eQTLs), transcriptomic resources, and gene-expression imputation studies have linked schizophrenia risk loci to genetically regulated expression, nominating putative risk genes and regulatory mechanisms across brain regions^3,6–8^. These studies support a regulatory model in which common genetic liability influences schizophrenia risk, at least in part, through distributed effects on brain gene expression and molecular pathways. However, how schizophrenia-associated regulatory variation is expressed at the level of individual patients and how such molecular liability maps onto macroscale brain network phenotypes remain incompletely understood.

Sex is a major source of heterogeneity in schizophrenia and related psychiatric disorders, with differences in incidence, age of onset, symptom profiles and cognitive burden^9–12^. More recently, Blokland et al. showed that SNP-based heritability estimates were substantially different between the sexes in schizophrenia, with the SNP showing top sex-interaction in schizophrenia being a cis-eQTL in several brain regions^13^. Recent postmortem transcriptomic work has further shown that sex shapes schizophrenia-associated molecular signatures, including sex-specific dysregulation and sex-interacting regulatory effects^14^. These findings suggest that schizophrenia-related molecular pathology may be partly sex-dependent. Yet direct evidence from living participants remains limited for sex differences in genetically-regulated transcriptomic alterations, i.e., sex-moderated regulatory liability, and how this can be translated into individualized brain functional network patterns that explain cognitive impairment and clinical symptom dimensions.

Functional magnetic resonance imaging (fMRI) studies have positioned schizophrenia as a disorder of distributed functional dysconnectivity^15–17^. Aberrant intrinsic network organization and functional network connectivity (FNC) have been observed across psychosis, supporting the dysconnectivity hypothesis that disrupted integration among large-scale brain networks may directly shape behavior and psychopathology^17–20^. In particular, abnormalities involving default-mode (DM), salience (SA), cerebellar (CB) and subcortical (SC) networks have been associated with cognitive deficits and positive and negative symptom dimensions in schizophrenia^21–24^. Imaging genetic studies further suggest that schizophrenia liability is reflected in functional connectivity: schizophrenia polygenic risk has been associated with altered connectivity across visual (VI), DM and frontoparietal networks, with relevance to cognitive ability^25,26^. In addition, sex-specific alterations in functional connectivity^27^ and sex-dependent associations between psychiatric polygenic risk and functional connectivity have also been reported^28^. Broader imaging-genetic analyses further indicate genetic overlap between multivariate functional connectivity measures and psychiatric disorders, including schizophrenia^29^. More recently, spatial dynamic subspace analyses showed that posterior DM/SA spatial functional network connectivity exhibits sex-specific schizophrenia alterations during a highly integrated brain state and is correlated with schizophrenia genetic risk^20^. Together, these studies indicate that schizophrenia-related dysconnectivity is genetically related and sex-modulated. However, most existing work has examined genetic liability, regulatory molecular variation, functional dysconnectivity, and sex influence as separate levels of analysis.

This work aims to investigate from an integrated view whether sex differences may exist in genetically-regulated transcriptomic variation and whether its relationship with functional connectivity may be moderated by sex in schizophrenia. Specifically, we hypothesized that schizophrenia-related regulatory liability and functional connectivity alterations would be captured by latent components within each modality, and that disease-associated components would exhibit coordinated multimodal relationships that may differ by sex. We independently decomposed genetically predicted gene-expression and FNC features. We first identified schizophrenia-associated gene-expression components that also showed sex differences, then systematically examined sex-dependent multimodal coupling between gene expression and FNC components, and characterized their molecular and network architecture.

## RESULTS

### Independent decomposition identifies multimodal gene-expression–FNC associations

We analyzed matched genotype-derived gene-expression and FNC data from 367 participants in the combined FBIRN^30^ and COBRE^31^ cohorts, including 160 neurotypical controls and 207 individuals with schizophrenia (mean age = 38.34 ± 11.92 years; 76 females/291 males). For each individual, brain tissue-specific eQTL-informed regulatory features were derived from genotype data using MetaXcan transcriptomic prediction models^32,33^, and FNC was constructed following the NeuroMark pipeline leveraging the NeuroMark 1.0 functional network templates^34^. Genetically regulated gene expression and FNC features were independently decomposed into 40 and 48 latent patterns or components, respectively.

We first examined whether sex differences might exist in schizophrenia-associated transcriptomic variation. We tested each gene-expression component for schizophrenia diagnosis effects and applied Bonferroni correction across all components, resulting in four components showing significant schizophrenia associations. We further examined whether any of these four components showed sex differences with Bonferroni correction, and found one schizophrenia-associated gene-expression component that also showed a marginally-significant sex difference (*p*_Bonf = 0.06).

We then focused on this gene-expression component to investigate how it might map to functional dysconnectivity and whether the multimodal association might differ by sex. To identify functional dysconnectivity, we tested each FNC component for schizophrenia diagnosis effects, and identified 23 components showing significant associations after applying Bonferroni correction. We then investigated the relationships between the target gene-expression component and these 23 FNC components showing schizophrenia-related dysconnectivity, which together defined 23 candidate multimodal component pairs. Among the 23 pairs, one pair showed a significant association between the gene-expression and FNC components after Bonferroni correction (*p*_Bonf = 1.72e-02) and was selected for detailed multimodal characterization, including evaluation of sex-interaction, as well as molecular and network interpretation. To further assess the generalizability of these findings, we also evaluated the gene-expression component and multimodal relationship for this pair of components in independent UK Biobank (UKB)^35,36^ data.

### Molecular architecture of schizophrenia-associated gene-expression component with sex difference

In the combined FBIRN and COBRE sample, subject loadings of the target gene-expression component were significantly lower in individuals with schizophrenia than in neurotypical controls (*β* = −0.30, *t* = −3.34, *p* = 9.37e-04, *p*_Bonf = 0.04, **Figure 1A**). In the meantime, this target component showed a marginally-significant sex difference, with the loadings being lower in males than females (*β* = 0.26, *t* = 2.42, *p* = 1.62e-02, *p*_Bonf = 0.06). We next characterized the molecular architecture underlying this component. Among 65,269 brain gene-by-tissue regulatory features, 3,490 exceeded the Bonferroni-corrected threshold (*p*_Bonf < 0.05, corresponding to |z| > 4.94, **Figure 1B**), representing 2,870 unique genes. Tissue-enrichment analysis revealed a pronounced cerebellar signature. Features contributing to the component were significantly enriched in the cerebellum (CER; fold enrichment = 1.63, *p*_Bonf = 0.0013, **Figure 1C**) and cerebellar hemisphere (CBH; fold enrichment = 1.39, *p*_Bonf = 0.0013, **Figure 1C**). Gene Ontology enrichment analysis^37^ further revealed convergence on intracellular transport and cellular structural processes. Prominent enriched terms included vesicle organization, membrane organization, protein transport, intercellular transport, endomembrane system organization, cilium organization, and microtubule-based transport (*p*_Bonf < 0.05, **Figure 1D**). Together, these findings indicate that there might be sex difference in the schizophrenia-associated gene-expression component which captures a distributed regulatory program preferentially represented in cerebellar tissues and centered on vesicle–membrane organization, intracellular trafficking, and microtubule-dependent transport.

**Figure 1.**
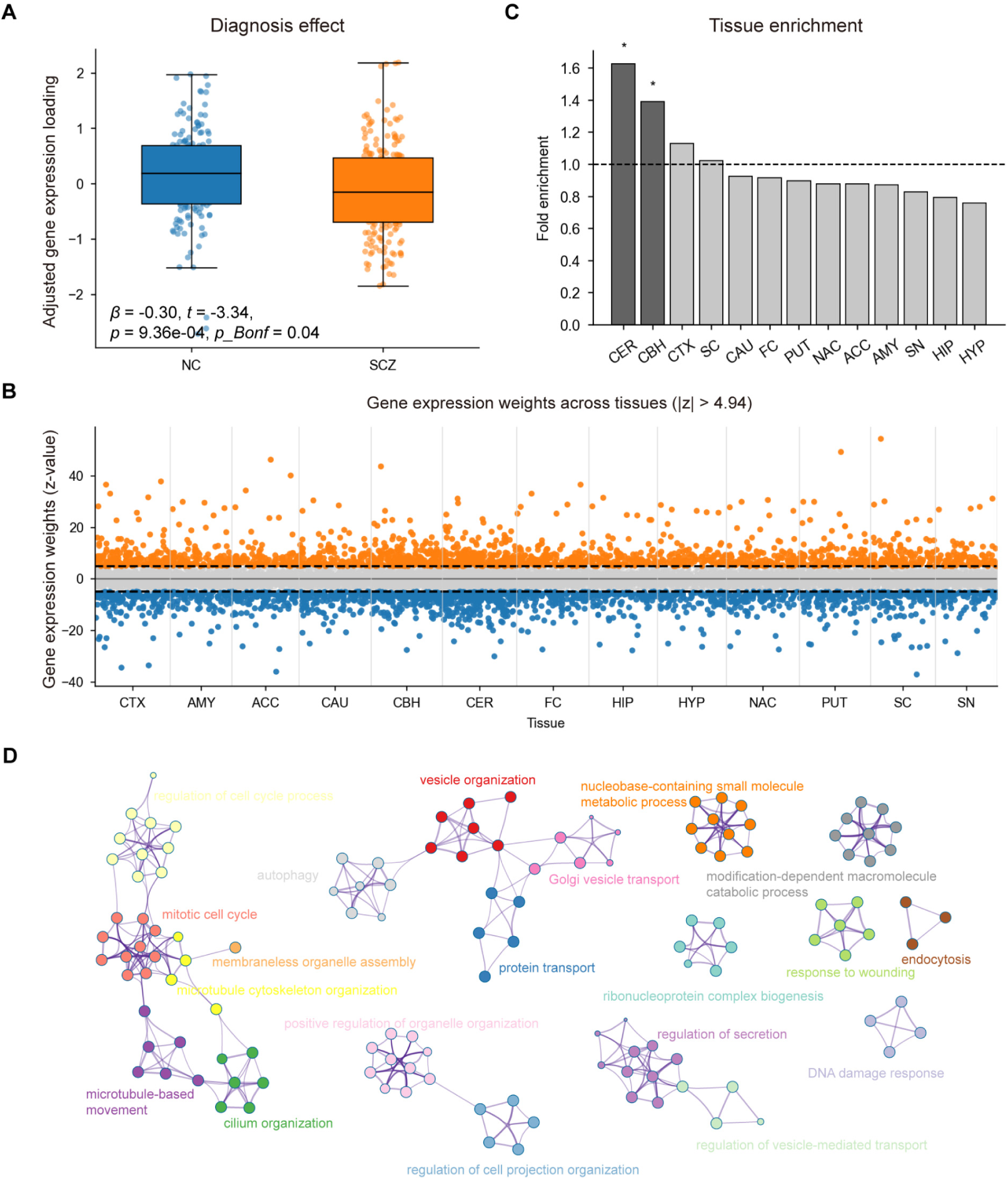
Molecular characterization of the schizophrenia-associated gene-expression pattern. **A**, Diagnosis effect on the selected gene-expression component in the combined FBIRN and COBRE sample. **B,** Gene-by-tissue weights of the selected component across 65,269 brain regulatory features; features exceeding the Bonferroni-corrected threshold are highlighted. **C,** Tissue enrichment of strongly contributing gene-expression features across 13 brain tissues. **D,** Gene Ontology enrichment of genes contributing to the selected expression pattern, highlighting intracellular transport, vesicle- and membrane-related, ciliary, and microtubule-associated processes.

### Sex-dependent multimodal association of the gene-expression component with FNC component

The FNC component paired with the target gene-expression component showed a strong diagnosis effect, with subject-level loadings significantly higher in individuals with schizophrenia than in controls (*β* = 0.49, *t* = 4.66, *p* = 4.44e-06, *p*_Bonf = 2.13e-04, **Figure 2A**). The corresponding connectivity pattern was distributed across multiple large-scale functional systems. Positive component weights were prominent within the cognitive-control (CC) domain and in cross-network connections involving subcortical (SC), default-mode (DM), and visual (VIS) systems. In contrast, negative weights were observed in connections including sensorimotor– cognitive-control (SM–CC) and cognitive-control–default-mode (CC–DM) interactions. A total of 56 FNC edges exceeded the Bonferroni-corrected component-weight threshold (*p*_Bonf < 0.05, corresponding to |z| > 4.13, **Figure 2B**).

**Figure 2.**
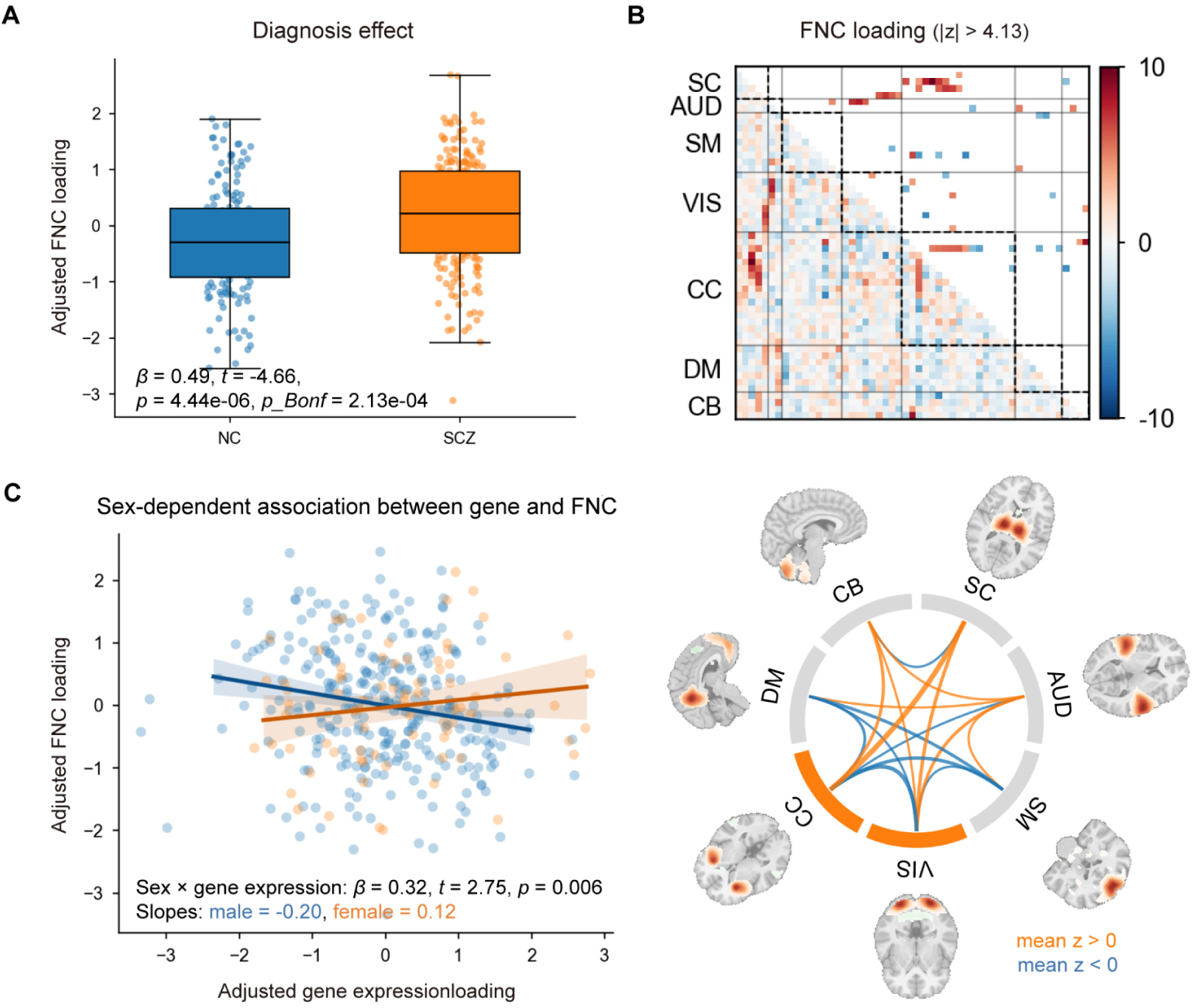
Schizophrenia-associated FNC pattern and its multimodal relationship with gene expression. **A**, Diagnosis effect on the selected FNC component in the combined FBIRN and COBRE sample. **B**, Connectivity-weight pattern of the selected FNC component across the 53 NeuroMark networks; edges exceeding the Bonferroni-corrected threshold are displayed in the network representation. **C**, Sex-dependent association between the selected gene-expression and FNC component loadings. Lines indicate fitted associations separately for males and females.

Regarding the gene-expression–FNC association, higher gene-expression loadings were associated with lower FNC loadings in the COBRE+FBIRN data (*β* = −0.20, *t* = −3.40, *p* = 7.47e-04, *p*_Bonf = 1.72e-02). We further examined whether this multimodal relationship differed by sex, and observed a significant sex-by-gene-expression interaction (*β* = 0.32, *t* = 2.75, *p* = 0.006, *p*_Bonf = 0.006, **Figure 2C**). The interaction reflected a negative gene-expression–FNC association in males and a weak positive association in females (male *r* = −0.20, female *r* = 0.12, **Figure 2C**), indicating that the multimodal relationship differed in both magnitude and direction across sex groups. This sex-moderated multimodal association suggests potential sex differences in the way how schizophrenia-related regulatory expression impacts functional network organization.

### UK Biobank evaluation

We evaluated the generalizability of these findings in independent UK Biobank samples. For the highlighted gene-expression component, we examined its projected loadings for sex and schizophrenia associations in 2,273 UKB participants, including 847 individuals with schizophrenia and 1,426 controls matched for age and sex. The schizophrenia diagnosis effect replicated strongly, with significantly lower component loadings in individuals with schizophrenia (*β* = −0.33, *t* = −7.75, *p* = 1.34e-14, *p*_Bonf = 5.36e-13, **Figure 3A**). The direction and magnitude of this effect consistently matched those observed in the COBRE+FBIRN discovery samples, providing strong independent support for the reproducibility of the schizophrenia-associated regulatory expression pattern. In the meantime, although this component showed no significant sex difference in UKB (*p* = 0.24), a significant sex-diagnosis interaction was noted (*p* = 1.15e-02), where a stronger sex difference was noted in the patient than control group.

**Figure 3.**
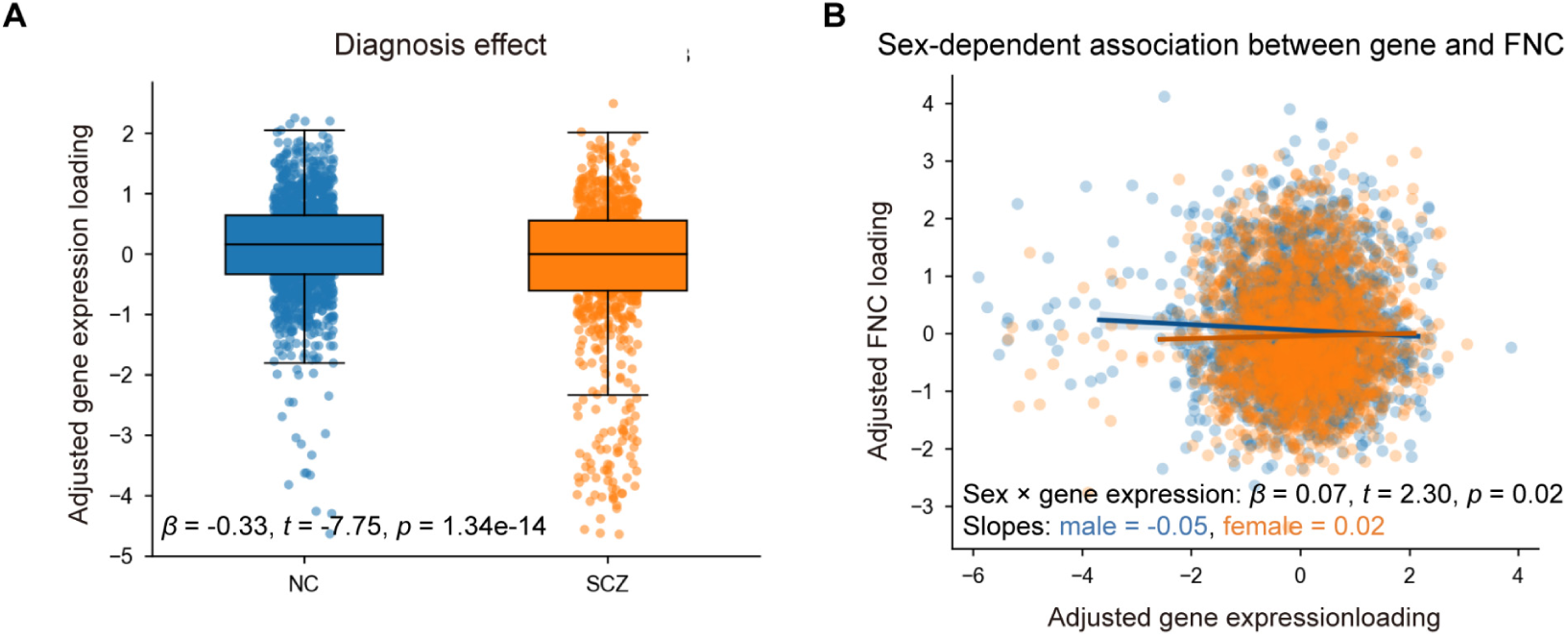
Independent evaluation in UK Biobank. **A**, Replication of the schizophrenia diagnosis effect on the selected gene-expression component in UK Biobank. **B**, Multimodal association between gene-expression and FNC component loadings stratified by sex in participants with matched genetic and imaging data. The male-negative and female-positive directional pattern was consistent with that observed in the discovery sample.

We next evaluated the multimodal relationship in 4,206 UK Biobank neurotypical participants with matched gene-expression and imaging data. The gene-expression– FNC association remained negative and significant (*β* = −0.05, *t* = −2.32, *p* = 0.02, **Figure 3A**), consistent with the direction observed in the discovery sample. The sex-by-gene-expression interaction was also directionally consistent (*β* = 0.07, *t* = 2.30, *p* = 0.02, **Figure 3B**) as in the discovery sample, with the association being negative in males (slope = −0.05) and weakly positive in females (slope = 0.02). Together, the UK Biobank analyses provided an independent replication of the schizophrenia-associated gene-expression pattern and its multimodal coupling with FNC and the associated sex-modulated pattern.

## DISCUSSION

In this study, we integrated genetically regulated brain gene-expression features with FNC to examine sex differences in schizophrenia-associated molecular variation and how it relates to large-scale brain network organization. By independently decomposing the two modalities, we identified a reproducible schizophrenia-related gene-expression pattern which showed a significant association with one functional dysconnectivity pattern. The molecular pattern was characterized by prominent cerebellar enrichment and intracellular transport- and membrane-related processes, whereas the associated FNC pattern involved distributed large-scale network dysconnectivity. Notably, the multimodal relationship also showed a consistent sex-dependent pattern across datasets, with a negative association in males and a positive association in females. Together, these findings support coordinated molecular and functional-network alterations in schizophrenia and suggest that their coupling may be moderated by sex.

Although postmortem transcriptomic studies provide direct evidence for molecular dysregulation in schizophrenia, comparable *in vivo* molecular-to-network evidence remains limited. Nevertheless, several lines of evidence suggest that schizophrenia-related functional dysconnectivity may be partially constrained by genetic and transcriptomic architecture. Topological patterns of gene expression correlations have been shown to recapitulate resting-state functional brain networks, and genetic variation in these gene sets has been linked to functional connectivity^38^. Imaging-genetic studies have further shown that schizophrenia polygenic risk is associated with altered functional connectivity across visual, default-mode and frontoparietal networks^25^. And recent multivariate genetic analyses demonstrate genetic overlap between functional connectivity and schizophrenia^29^. Together with evidence that brain organization is coupled to underlying cellular and molecular architecture^39–41^, these findings provide a biological basis for examining whether schizophrenia-associated regulatory and functional abnormalities converge at the individual level.

Multimodal fusion approaches can capture both shared and modality-specific abnormalities across biological systems^42–44^. This is particularly relevant to schizophrenia, where molecular and connectomic alterations are distributed, modest in magnitude, and heterogeneous across individuals. We therefore used an ICA-based framework to identify latent schizophrenia-related components independently within gene-expression and FNC data, and then further evaluated multimodal associations. This strategy preserves modality-specific organization and allows multimodal relationships to emerge from independently defined disease-related patterns.

Within this framework, we identified a pair of components where the loading of the gene-expression pattern was significantly associated with the loading of the FNC pattern. Importantly, this relationship showed a significant sex interaction, with a stronger negative association in males and a weaker positive association in females, and the same directional pattern was observed in normal participants of UKB. This is biologically plausible given the documented evidence, including reports of sex difference in SNP-based heritability^13^, substantial effects of sex on gene expression and genetic regulation across human tissues^45^, recent BrainSeq finding of sex-specific schizophrenia-associated transcriptional dysregulation and sex-interacting eQTL effects^14^, as well as sex-dependent relationships between schizophrenia polygenic liability and cognitive impairment^12^. Although a smaller effect size was noted in the UKB normal participants, the consistency across datasets suggests that sex may modify how schizophrenia-related regulatory variation is expressed at the level of large-scale brain networks, and that this sex-moderated association likely holds in both control and schizophrenia population.

The molecular architecture of the expression pattern further points to intracellular trafficking and cytoskeletal processes that have been associated with schizophrenia. Postmortem transcriptomic studies have reported abnormalities in Golgi function, vesicular transport, membrane association, and presynaptic vesicle trafficking^46,47^. Accumulated evidence from genetic and transcriptomic analyses has implicated endosomal trafficking and microtubule-dependent transport pathways^48^. Cilia-related programs may represent an additional convergent mechanism, as schizophrenia-associated genes have been linked to alterations in primary-cilium organization and intraflagellar transport^49^. Notably, the strong cerebellar enrichment observed in this study is also aligned with previous postmortem evidence of cerebellar abnormalities involving vesicular transport and membrane-associated processes^45^. Together, these findings suggest that the molecular programs implicated by the highlighted gene-expression pattern echo cellular pathways previously linked to schizophrenia, particularly those governing intracellular trafficking, cytoskeletal organization, and membrane-associated processes.

The paired FNC pattern showed a complementary systems-level organization, with prominent connections involving cerebellar, cortical, and subcortical systems. This distributed configuration is consistent with the broader dysconnectivity framework of schizophrenia^15,16,50^, and also aligned with the cognitive dysmetria model emphasizing disrupted cortical–subcortical–cerebellar coordination^51,52^. Together, the molecular and FNC findings suggest that cerebellar and distributed network abnormalities may represent coordinated manifestations of schizophrenia-related biology across scales. It remains to be elucidated how the gene-expression and FNC components further relate to clinical dimensions including cognitive impairment^24,53^ and psychotic symptoms^54,55^. The current findings should be interpreted in light of several limitations. First, eQTL-informed gene-expression features are genetically predicted regulatory proxies rather than direct measurements of brain gene expression and cannot fully capture cell-type-specific, developmental, environmental, or state-dependent transcriptional processes. Second, the highlighted gene-expression component presented different forms of sex dependency in the COBRE+FIBRN and UKB data, with the former showing a marginally-significant sex difference in component loadings and the latter showing a significant sex-diagnosis interaction. Additional data is needed to verify the observed sex effect. Third, a comprehensive characterization is warranted for the effects of medication exposure, illness duration, symptom state, cohort heterogeneity on the multimodal associations. Finally, sex was modeled as a biological variable, whereas gender-related psychosocial factors were not assessed. Future work should contextualize these relationships in larger, more sex-balanced, longitudinal, ancestrally diverse datasets of psychotic populations for independent multimodal replication.

In summary, this study identifies coordinated schizophrenia-associated molecular and functional-network patterns across biological scales. The observed multimodal coupling and its consistent sex-dependent direction across datasets highlight a potential sex-moderated regulatory-to-connectome pathway that warrants further independent validation and contextualization.

## MATERIALS AND METHODS

### Participants

We analyzed participants from four cohorts: FBIRN^30^, COBRE^31^, and UK Biobank^35,36^. The primary multimodal discovery sample consisted of 367 participants from the combined FBIRN and COBRE cohorts with both genotype-derived gene-expression and resting-state fMRI data, including 160 neurotypical controls and 207 individuals with schizophrenia. For independent evaluation, the UKB analyses used two subsets according to data availability. The gene-expression sex- and diagnosis-association analysis included 2,273 participants, comprising 847 individuals with schizophrenia and 1,426 normal participants as controls, whereas the multimodal analysis included 4,206 normal participants with matched genotype-derived gene-expression and imaging data. Demographic characteristics are summarized in **Table 1**. All datasets were collected under protocols approved by the relevant local institutional review boards, and all participants provided written informed consent according to the procedures of the original studies. The UKB data used in our work were obtained under application 34175.

**Table 1.** Demographic characteristics of the genetic and fMRI samples.

|  | Dataset | Samples | Diagnosis(NC/SCZ) | Age (mean±std) | Sex (F/M) |
| --- | --- | --- | --- | --- | --- |
| SNP | FBIRN+COBRE | 367 | 160/207 | 38.34±11.92 | 76/291 |
|  | UKB | 2273 | 1426/847 | 55.98±7.49 | 823/1450 |
| SNP-fMRI | FBIRN+COBRE | 367 | 160/207 | 38.34±11.92 | 76/291 |
|  | UKB | 4206 | 4206/0 | 56.22±7.38 | 2133/2062 |

### Genotype processing

For COBRE and FBIRN, the genotypes were imputed following the ENIGMA protocol and went through standard quality control, as described in our previous studies^56,57^. For UKB, the released imputed genotype data contained approximately 96 million variants, with the genotyping and imputation protocols described previously^36^. Variants with minor allele frequency below 0.01 were excluded^58^. For cross-cohort harmonization, UKB was used as the reference SNP set when merging genotype data across cohorts. SNPs from FBIRN and COBRE were aligned to the UK Biobank SNP set. Individual relatedness (identify-by-descent) was estimated using PLINK^59^; for groups of participants who were second-degree relatives or closer, one individual was retained for downstream analysis. Genetic principal components were computed from genome-wide SNP data, and the top 10 principal components were used as covariates for population stratification.

### Brain eQTL-informed regulatory features

Brain tissue-specific genetically regulated expression features were generated using MetaXcan^32,33^ transcriptomic prediction models. We focused on 13 GTEx^60^ brain tissues: amygdala, anterior cingulate cortex BA24, caudate basal ganglia, cerebellar hemisphere, cerebellum, cortex, frontal cortex BA9, hippocampus, hypothalamus, nucleus accumbens basal ganglia, putamen basal ganglia, spinal cord cervical C1, and substantia nigra. This procedure yielded 65,269 gene-by-tissue expression features for 12,268 genes. Each feature represented the genetically predicted expression of a gene in a specific brain tissue.

### Imaging preprocessing and FNC construction

All preprocessing was performed using the Statistical Parametric Mapping (SPM12, http://www.fil.ion.ucl.ac.uk/spm/) toolboxes within the MATLAB 2024b environment. The preprocessing included rigid-body motion correction, slice-timing correction, spatial normalization to the MNI template using an EPI-based approach, resampling to 3 × 3 × 3 mm³ isotropic voxels, and spatial smoothing with a 6-mm FWHM Gaussian kernel. Preprocessed fMRI data were decomposed into 53 subject-specific intrinsic connectivity networks and their corresponding time courses using Multivariate Objective Optimization Independent Component Analysis with Reference (MOO-ICAR)^34^, implemented in the GIFT toolbox (http://trendscenter.org/software/gift and http://trendscenter.org/data) with NeuroMark 1.0 functional templates^34^. Time courses were denoised using detrending to remove linear, quadratic, and cubic trends; outlier detection and removal; nuisance regression of six rigid-body motion parameters and their temporal derivatives; and temporal band-pass filtering from 0.01 to 0.15 Hz^27^. Subject-level FNC matrices were computed as pairwise Pearson correlations among the 53 denoised network time courses, resulting in 1,378 unique FNC features per participant. We further regressed out site effects (dummy-coded) separately in the discovery COBRE+FBIRN and replication UKB data for each FNC feature and the residualized FNC features were used in the subsequent analyses.

### Independent decomposition of gene-expression and FNC features

To identify distributed latent components within each modality while preserving modality-specific structure, genetically regulated gene-expression and FNC features were decomposed separately using independent component analysis (ICA)^61,62^. Before ICA, the principal component analysis (PCA) with reference method^63^ was utilized for dimension reduction and whitening, where diagnosis served as the reference to retain schizophrenia-related variance in the analysis. The gene-expression data were represented by 40 independent components and the FNC data by 48 independent components, yielding, for each component, a feature-weight pattern and a subject-level loading. The model orders 40 and 48 were determined to capture 99% of diagnosis-related variance in the original data. The two modalities were decomposed separately rather than using gene-expression information to guide FNC decomposition, which was to avoid overfitting given the limited discovery sample size of COBRE+FBIRN. This design enabled subsequent multimodal relationships to be evaluated between independently derived latent components.

### Identification of schizophrenia-associated gene-expression components with sex differences

Subject-level component loadings were tested for schizophrenia diagnosis effects separately within each modality. For gene-expression components, loadings were standardized and modeled as a function of sex, diagnosis, and the first 10 genetic principal components (as covariates for population stratification). A secondary model additionally included the sex-by-diagnosis interaction. Thus, the models were

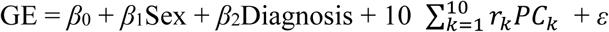

and

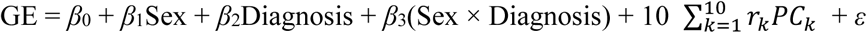

GE stands for gene expression. Diagnosis effect *p*-values were Bonferroni-corrected across all components. Components with *p*_Bonf < 0.05 were considered schizophrenia-associated. We further examined whether there might be significant sex or sex-diagnosis interaction effects (*p*_Bonf < 0.05) within the schizophrenia-associated components. This procedure identified one schizophrenia-associated gene-expression component that also showed a significant sex difference.

### Identification of multimodal association

To identify FNC components showing schizophrenia-related dysconnectivity, standardized subject loadings were modeled as a function of standardized age, sex, and diagnosis, with a secondary model including the sex-by-diagnosis interaction (site effects already corrected at the feature level):

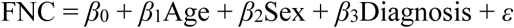

and

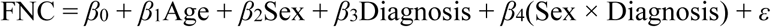

Similarly, diagnosis effect *p*-values were Bonferroni-corrected across all components, which led to 23 components with *p*_Bonf < 0.05 indicating functional dysconnectivity. Notably, we did not observe any significant sex differences among these 23 FNC components.

Multimodal associations were then evaluated among the identified schizophrenia-associated gene-expression component with sex difference and the FNC components showing schizophrenia-associated dysconnectivity. To minimize confounding by population structure, each gene-expression component loading was first residualized for an intercept and the first 10 genetic principal components and subsequently standardized. FNC component loadings and age were also standardized. For each candidate pair in the COBRE+FBIRN discovery sample, the following model was fitted:

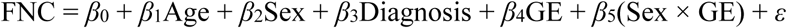

This model exactly reflects the current implementation, where the expression main effect tested multimodal coupling between the two components, whereas the sex-by-GE term tested whether this coupling differed between males and females. We then asked among those biologically-associated gene-expression and FNC components, whether any gene-expression–FNC relationship might be sex-dependent. Consequently, we first identified the components pairs with significant gene-expression main effects after correcting for 23 candidate pairs using Bonferroni correction. We then examined whether any sex-by-GE effect was significant with Bonferroni correction within the pairs where a significant main effect was observed.

### Molecular characterization of the selected gene-expression component

To characterize the molecular architecture of the selected component, component feature weights across the 65,269 gene-by-tissue regulatory features were standardized to *z* values. Two-sided probabilities were calculated from the standard normal distribution and Bonferroni-corrected across all 65,269 features. Features with *p*_Bonf < 0.05 were retained for downstream characterization. Tissue enrichment was assessed by comparing the representation of significant features within each of the 13 brain tissues against their background representation among all gene-by-tissue features. Statistical significance was evaluated using permutation testing followed by Bonferroni correction across tissues. Genes represented by strongly contributing features were further subjected to Gene Ontology enrichment analysis using Metascape^37^ to identify biological processes associated with the component.

### Characterization of the selected FNC component

FNC edge weights of the selected component were standardized to *z* values. Two-sided probabilities were calculated under the standard normal distribution and Bonferroni-corrected across all 1,378 FNC edges. Edges with *p*_Bonf < 0.05 were retained to characterize the network architecture of the component. Because the sign of an ICA component is arbitrary, positive and negative edge weights were interpreted as opposing poles of the latent FNC pattern rather than direct evidence of schizophrenia-related hyperconnectivity or hypoconnectivity.

### UK Biobank evaluation

The identified gene-expression and FNC components were projected onto the UKB data, resulting in subject-level loadings that reflected how the component patterns identified in the discovery samples were loaded on individuals of UKB. This projection strategy allows flexible individual-level estimates in unseen data rather than requiring re-estimation. The schizophrenia association of the selected gene-expression component was evaluated independently in 2,273 UKB participants with schizophrenia diagnosis information. As in the discovery analysis, standardized component loading was modeled as a function of sex, diagnosis, and the first 10 genetic principal components, and a nominal *p* < 0.05 along with consistent direction of effect was considered as successful replication, given that the analysis targeted only one component pair without involving multiple hypotheses. Multimodal generalizability was subsequently evaluated in 4,206 UKB neurotypical participants with matched gene-expression and imaging data. Gene-expression component loadings were residualized for the first 10 genetic principal components and standardized, and the corresponding FNC loadings and age were standardized. Because only a very limited number of individuals with schizophrenia had both imaging and genotype data available, the UKB multimodal analysis was restricted to neurotypical participants. Consequently, diagnostic status was not included as a variable in this model.

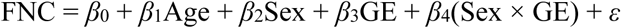

Similarly, for the expression main effect and sex-by-expression interaction effect, a nominal p < 0.05 along with consistent direction of effect was considered successful replication.

## DATA AVAILABILITY

The FBIRN and COBRE datasets require application or controlled access and can be obtained from the repository (https://coinstac.org/) or investigators. The UK Biobank dataset used in this study are publicly available through application (https://www.ukbiobank.ac.uk/). NeuroMark 1.0 template of intrinsic functional networks (http://trendscenter.org/software/gift).

## CODE AVAILABILITY

Software used includes SPM12 (http://www.fil.ion.ucl.ac.uk/spm/) and the GIFT toolbox (http://trendscenter.org/software/gift).

## ACKNOWLEDGEMENT

The authors thank all the volunteers for their participation in the study and the anonymous reviewers for their insightful comments and suggestions.

